# Prevalence and Penetrance of Heritable Retinoblastoma in Two Adult Population Cohorts: Implications for genomic newborn screening

**DOI:** 10.64898/2025.12.16.25342350

**Authors:** Isabella-Anna Lazaridi, Timothy Hall, Helen Hanson, James Fasham, Emma L. Baple, Michael N. Weedon, Caroline F. Wright, Leigh Jackson

## Abstract

Retinoblastoma (Rb) is a rare childhood eye cancer. Almost half of cases are heritable, associated with germline *RB1* pathogenic variants that pre-dispose to Rb and extraocular cancers. This study aimed to investigate the prevalence and penetrance of *RB1*-heritable Rb in two adult population cohorts. We screened participants with whole genome sequencing in the UK Biobank (UKB) (n=490,413) and All of Us (AoU) (n=317,964) cohorts for predicted loss-of-function (pLoF) and/or ClinVar pathogenic/likely pathogenic *RB1* variants. Electronic health records and questionnaires were used to screen participants for Rb-associated features. In the UKB we generated a stringent and permissive phenotype category, ranging from Rb to Rb-associated extraocular cancers; in AoU we included participants with Rb or ocular cancer. A total of 22 pathogenic pLoF *RB1* variants were detected in the UKB (n=12) and AoU (n=13) participants. In the UKB, only 25.0% (3/12) of variant carriers reported developing Rb by the age of 60, increasing to 50.0% when including ocular/extraocular cancers. Similarly, 30.8% (4/13) AoU participants developed Rb and/or ocular cancer by the age of 60. Overall, this results in a combined penetrance estimate of 28.0% (7/25). We found 21 and 28 individuals with Rb in the UKB and AoU, respectively, which is within published prevalence estimates, suggesting these cohorts are not depleted of Rb cases. Notably, the penetrance of pathogenic *RB1* variants in >800,000 clinically unselected adults was substantially lower than the near complete penetrance reported in clinical cohorts. This has important implications for counselling families following a positive newborn screening result.

## Introduction

Public newborn screening programmes have been available worldwide for many years for a limited set of conditions (1,2). Recently there has been increasing interest in using genome sequencing to enable a genotype-first approach to screening for many more conditions, aiming to test pre-symptomatic newborns for preventable and/or treatable rare genetic disorders (2). With decreasing costs and advancements in the latest sequencing technologies, genomic newborn screening (gNBS) programmes are now a feasible option to consider introducing into public health programmes (2,3). While many benefits have been proposed, it is essential to understand the challenges associated with implementing pre-symptomatic screening for rare genetic variants in asymptomatic newborns. Genotype-first screening approaches can be ambiguous, especially as variant penetrance and age of onset can vary between clinically ascertained and population cohorts (4). This can make variant interpretation challenging if there is uncertainty around whether the newborn will go on to develop the condition. With >4,000 different genes being considered for inclusion across different gNBS studies, we need a better understanding of the penetrance of these pathogenic variants in population cohorts (5)

The *RB1* and heritable retinoblastoma (hRb) gene-disease pair is being assessed in at least 21 gNBS research studies currently in progress, including the Generation Study (5). Retinoblastoma (Rb) is a rare malignant neoplasm of the retina, with an incidence of 1 in 15,000-20,000 live births, or a 1-9 in 100,000 lifetime prevalence in Europe (6,7). Almost all cases are diagnosed under five-years of age, most presenting before 12 months (6). Most cases are caused by loss-of-function (LoF) of the *RB1* gene, and approximately 40-50% are inherited (8,9). hRb is an autosomal dominant cancer pre-disposition syndrome, where a germline *RB1* variant substantially increases the risk of developing Rb (8–11). Although Rb has a near 100% five-year survival rate (13), individuals with hRb are at risk of undergoing uniocular or binocular enucleation, and at increased risk of developing associated extraocular cancers, which can occur as early as 10 years of age (8–11).

In the UK, the National Health Service (NHS) offers genetic testing for individuals with Rb to detect germline *RB1* variants and inform ongoing management and at-risk family members (14). For those with a positive family history, genetic testing and appropriate screening measures can be initiated prior to symptom onset (15,16). Without a family history, genetic testing for germline *RB1* variants would only currently occur after a clinical diagnosis has been made (17). Current guidelines recommend frequent surveillance from birth, followed by regular screening until 5-7 years of age (9,15,16). This involves a dilated fundus examination requiring multiple hospital visits, with examinations under anaesthesia (EUA) between six months to three years of age, if adequate views cannot be obtained when the child is awake (9,15,16).

hRb is considered a highly penetrant condition, with reports in the literature of near complete penetrance (>90%) (7,8,18). Although low penetrance variants have been reported, these are usually whole gene deletions encompassing the *MED4* gene, missense, promoter and certain splicing variants with in-frame exon skipping (19). Highly penetrant variants, usually nonsense, frameshift and splicing variants, will often result in premature termination codons (PTCs) and induce nonsense mediated decay (NMD) of the transcript leading to diminished or absent levels of protein (19,20). The high reported *RB1* variant penetrance has been derived from clinically ascertained cohorts (19–21) and thus is likely to be an over-estimate with respect to population penetrance (4,22,23). This has created a lack of understanding of the penetrance of these variants when ascertained in the general population, which is essential for informing newborn screening (4,24). Considering one of the key inclusion criteria for gNBS programmes is that a condition needs to be highly penetrant, a better understanding of the population *RB1* variant penetrance is required to inform gNBS guidelines, aid variant interpretation, and provide families with accurate risk estimates.

In this study, we aimed to assess the prevalence and penetrance of *RB1* variants using the UK Biobank (UKB) and All of Us (AoU) research programme, two adult population-based cohorts, to improve our understanding of *RB1* variant risk in the context of screening.

## Methods

### Cohorts

#### UK Biobank

The UKB is a large-scale, population-based cohort made up of 502,355 participants >40 years of age at recruitment, between 2006-2010 across the UK (25). At registration, participants provided biospecimens and completed interviews and questionnaires, including information on self-reported medical conditions and operations (25). Electronic Health Records (EHRs), including hospital episode statistics (HES) and cancer registry data were available for all participants. Short-read whole genome sequencing (WGS) and exome sequencing data was available for 490,413 and 454,712 participants, respectively. Use of this data was approved by the UK Biobank Research and Ethics Committee, under the approved project “Understanding the role of rare and common genetic variation in human phenotypes”, application number 103356.

#### All of Us

The AoU research programme is a large-scale population cohort recruiting individuals ≥18 years of age across the USA since 2018, currently made up of 633,540 participants. Registration is completed through an AoU-approved healthcare provider or online. Upon recruitment and consent, completion of a baseline health questionnaire is required. Participants can opt in to provide additional information, including questionnaires, EHRs and biospecimens (26). Short-read WGS was available for 414,830 participants, of which 317,964 also had EHR records. For this study we used the AoU research programme-controlled tier v8 dataset, available to authorised users on the Researcher Workbench. We have been granted an exception to the Data and Statistics Dissemination Policy from the AoU Research Access Board, allowing us to report participant counts <20.

### Genetic variant curation

The UKB and AoU sequencing methods are described in more detailed by the UKB Research Consortium (27,28) and AoU Research Programme Genomics Investigators (29). For both the UKB and AoU, variants in genome build GRCh38 were annotated using Ensembl’s Variant Effect Predictor (VEP) (30). Variant curation was consistent across both cohorts. Only variants in the *RB1* MANE select transcript (ENST00000267163.6/NM_000321.3) were considered. Extensive quality control was completed to ensure only putatively pathogenic variants were retained (**Figure 1**). We included variants predicted to cause LoF (pLoF) (annotated VEP ‘high-impact’) and considered ‘high-confidence’ according to the Loss-Of-Function Transcript Effect Estimator (LOFTEE) (31). This included frameshift, splice donor/acceptor and stop gain variants. In addition to pLOFs, we included any variants previously classified as Pathogenic and/or Likely Pathogenic (P/LP) at a 2* level or above, in the ClinVar database (accessed Nov-2024) (32). Potentially qualifying variants were manually checked on the ClinVar website. We further inspected variants using the Integrative Genomics Viewer (IGV) to assess the validity and genotype quality of each variant call. Specifically, calls with a read depth <3 or variant allele fraction (VAF) <27% were excluded. This VAF cut-off was used as previous literature suggested it had the optimal discriminatory power to distinguish germline from somatic mosaic variants (33). Exome sequencing data available for the same participants in the UKB was also used for variants with ambiguous or low quality WGS data.

**Figure 1:**
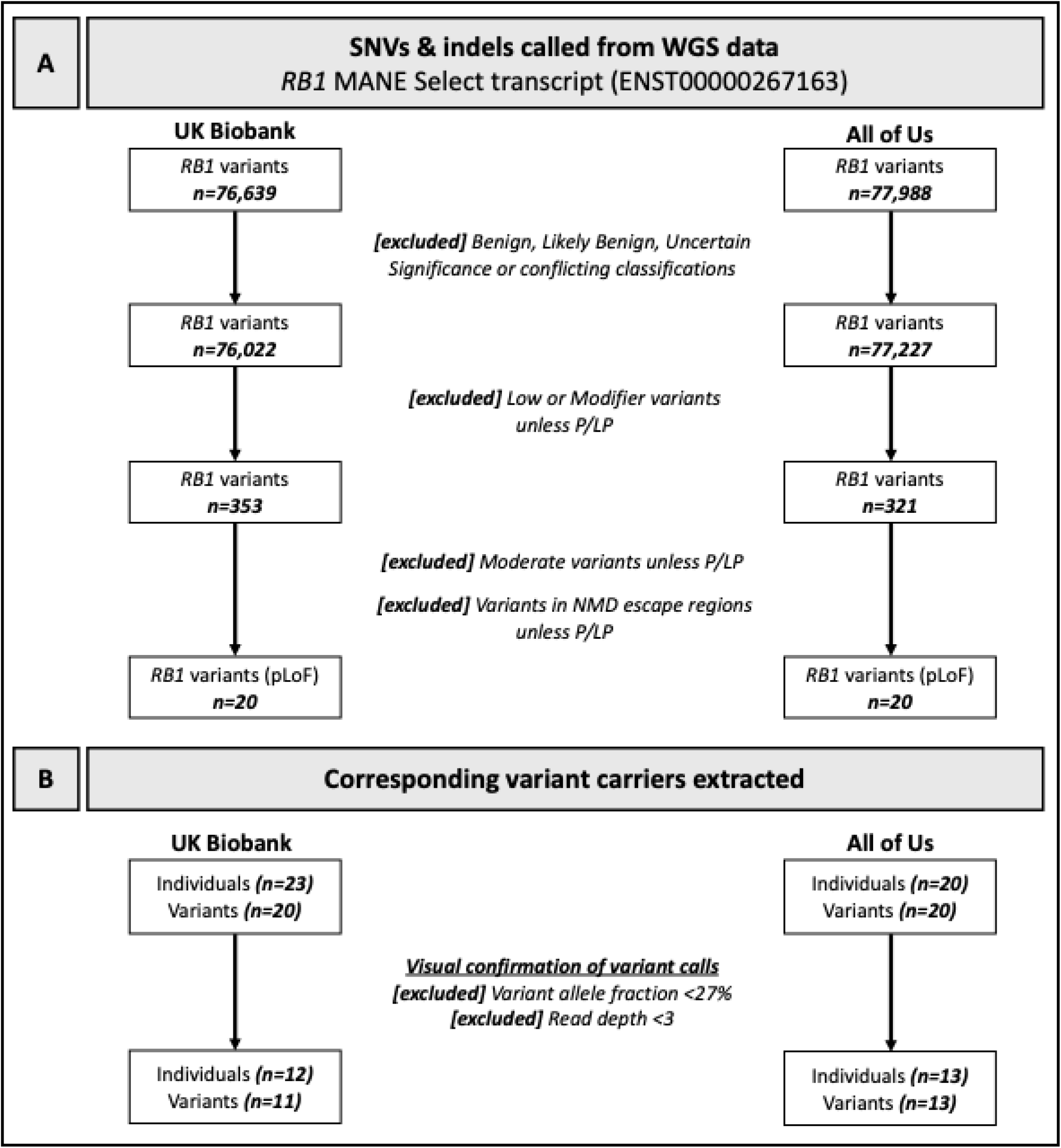
Flow chart of variant and genotype curation. Participants in the UK Biobank (n=490,413) and All of Us (n=317,964) with whole genome sequencing (WGS) data were screened for single nucleotide variants (SNVs) and small insertions or deletions (indels) in the *RB1* gene in. (A) Initially benign variation was excluded, followed by low, modifier or moderate impact variants, unless pathogenic (P) or likely pathogenic (LP) at 2* level or above and associated with retinoblastoma. If required, predicted loss of function (pLoF) variants in nonsense mediated decay escape regions were excluded, unless classified as P/LP. (B) Participants carrying any of the high confidence pLoF *RB1* variants were extracted. Each variant call was visually assessed, and participants were excluded based on the variant call quality, read depth or variant allele fraction.

### Classifying phenotype criteria

#### UK Biobank

We used a combination of cancer registry, hospital episode statistics (HES) records, and self-reported medical information to screen participants who had reported codes consistent with hRb. Associated phenotypes were mapped to corresponding international classification of disease codes (ICD-9/10). For associated procedures or operations, Office of Population Censuses and Surveys codes (OPCS-4) were used. We used self-reported medical information to detect those who may have been diagnosed prior to the earliest HES or cancer registry record available (34), specifically for Rb and/or eye cancers. We took the youngest age of diagnosis across all three datasets. We used two phenotype criteria, stringent and permissive, to encompass all individuals with hRb. Stringent phenotype criteria were consistent with a prior report of Rb or the associated ICD-9/10 code (1905/C69.2). Permissive phenotype criteria included any eye cancer, associated non-ocular cancers and procedures. A list of the ICD-9/10, OPCS-4 codes and self-reported cancers used for stringent and permissive criteria are outlined in **Table S1**.

#### All of Us

We combined participant EHRs and, where available, survey responses to classify Rb and eye cancer in the AoU cohort. EHRs were available from the 1980s onwards, which could exclude older individuals diagnosed prior to this date (35). Within survey responses, participants report their age of diagnosis within five categories (child, adolescent, adult, older adult and elderly). As the AoU research programme does not have dedicated cancer registry data, we used a broader set of phenotype codes to extract participants with features of hRb. We used SNOMED CT codes that best mapped to Rb and/or ocular cancer (**Table S2**). This approach was in line with literature studying cancer prevalence in AoU (36). We also used the “Personal and Family Health History” survey, which includes information on self-reported eye-cancer diagnoses (**Table S2**).

### Data analysis and statistical testing

Fisher’s exact test was used to quantify the association of binary traits in *RB1* pathogenic variant carriers and non-carriers. For tests involving multiple comparisons, a Bonferroni correction was applied. Penetrance was reported as the proportion of *RB1* pathogenic variant carriers matching our phenotype criteria. Kaplan-Meier survival analysis was completed to determine the age-related penetrance of Rb in individuals with germline *RB1* variants in the UKB. Failure was set as the age when individuals received a diagnosis for any feature in each phenotype category. Age at risk started from birth and ended at the most recent EHR data update in the UKB (2023) (37). Data handling and statistical testing were completed on the UKB DNAnexus platform and AoU researcher workbench, which includes the cohort builder, dataset builder and cloud workspace, using RStudio (v4.4.0) and Jupyter notebook (v6.5.4) with python (v3.10.16).

## Results

### Carriers of *RB1* pathogenic variants in the UK Biobank exhibit reduced penetrance compared to previously reported estimates

Screening WGS data from 490,413 UKB participants identified 11 high-confidence (likely) pathogenic or pLoF *RB1* variants in 12 individuals **(Table 1).** One variant (ENST00000267163.6:c.1666C>T (p.Arg556Ter)), was called as homozygous in one participant, with a VAF >90%, however confirmation in exome sequencing data revealed a lower VAF (71%) indicative of a heterozygous germline variant and consistent with the monoallelic requirement of hRb. All other individuals were heterozygous with 27% ≤ VAF ≤80% in WGS data (ref). Almost all variants had been previously submitted to the ClinVar database and asserted as P/LP with varying levels of evidence at the time of this study (**Table 1**). The genomic position of each variant against 2* P/LP ClinVar variants is visualised in **Figure 2**. UKB participants were also screened for diagnostic codes associated with each phenotype category, specifically stringent (Rb, n=21) and permissive (any ocular and Rb-associated extraocular cancers, n=49,544). We found a significant enrichment of both stringent and permissive phenotypes in *RB1* variant carriers; p=2.98x10^-11^ (OR:10,922) and p=1.07x10^-4^ (OR:12.5) respectively. Of the 12 *RB1* pathogenic variant carriers, only 25.0% (3/12) had diagnostic codes or self-reported conditions matching the stringent phenotype criteria by 60 years of age. Relaxing our phenotype criteria to include any ocular and Rb-associated non-ocular cancers increased the penetrance to 50.0% (6/12) by 60 years of age. One qualifying splicing variant, 13:48456350:GT:G (ENST00000267163.6:c.1960+3del) (Table 1), had a low SpliceAI score (<0.20), likely because the deleted ‘T’ was replaced by the same base. It was absent from gnomAD (non-UKB v4.1.0), and other variants at this position have been classified as 2* P/LP. Both carriers also carried a low-impact, 1*LP splicing variant (ENST00000267163.6:c.1960+5G>A) in cis. Although this variant was included in the main analysis, excluding these carriers would not significantly alter penetrance estimates, only increasing to 30.0% (3/10) and 60.0% (6/10) for stringent and permissive phenotypes, respectively.

**Figure 2:**
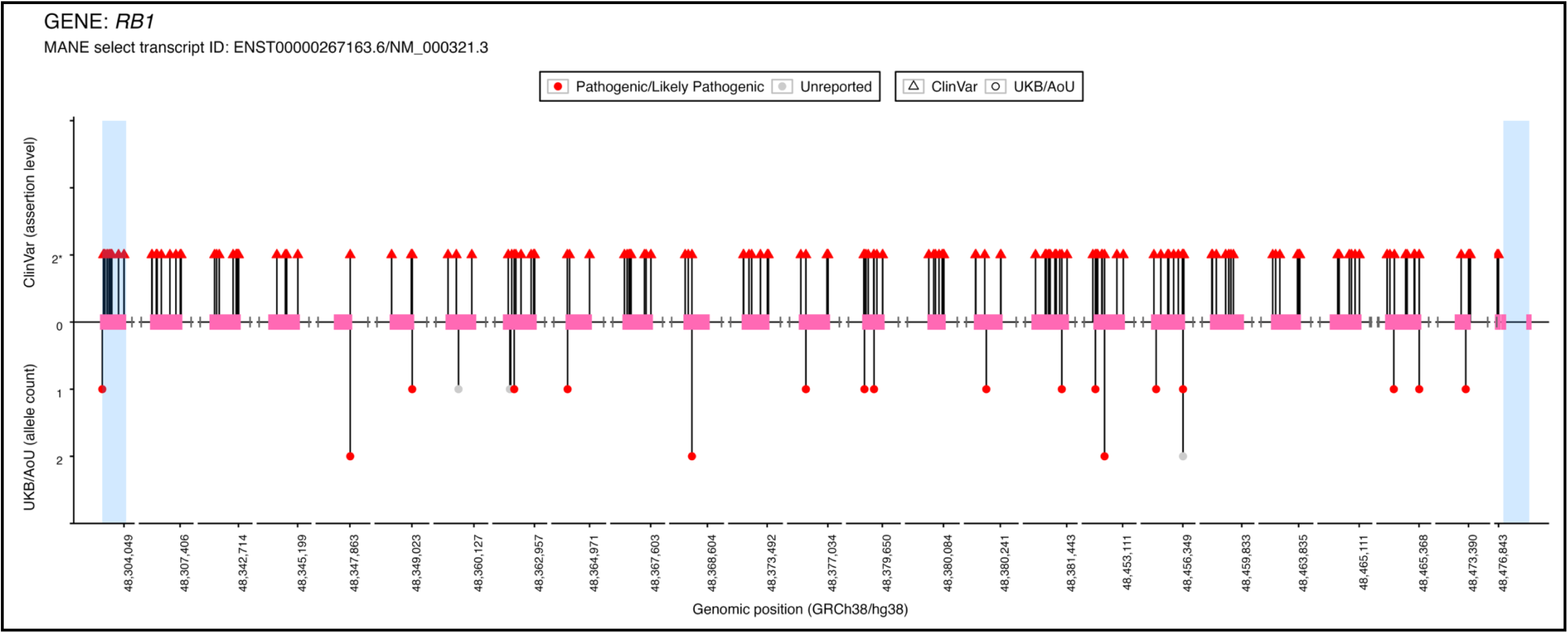
Lollipop plot of detected *RB1* variants in the UK Biobank (UKB) and All of Us (AoU) compared against ClinVar Pathogenic/Likely Pathogenic *RB1* variants. Plot of the 22 pathogenic *RB1* variants detected from the UKB and AoU cohorts compared to 219 known 2* Pathogenic/Likely Pathogenic *RB1* variants present in the ClinVar (accessed Nov 2024). ClinVar variants are depicted in the upper region of the plot. UKB/AoU variants are in the lower region of the plot, with the allele count shown on the y axis. Exons 1-27 for the *RB1* MANE Select transcript (ENST00000267163.6) are depicted by the pink boxes. The axis between exons has been split to remove long intronic regions. Regions highlighted in blue depict nonsense mediated decay escape regions.

**Table 1.**
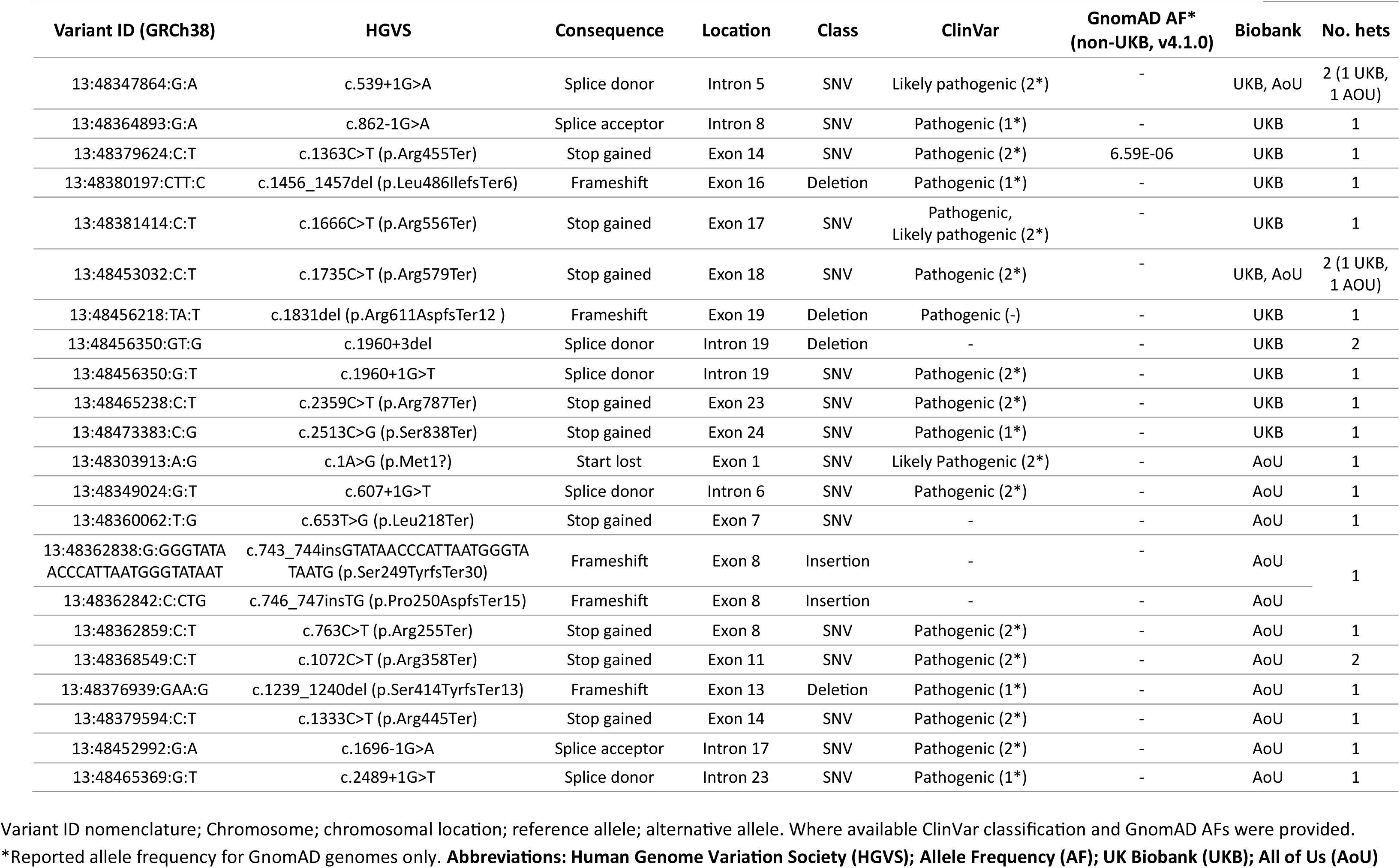
*RB1* variants (MANE Select-ENST00000267163.6) detected from whole genome sequencing in UK Biobank and All of Us cohorts.

We also used a Kaplan-Meier survival analysis to assess the age-related penetrance of *RB1* pathogenic variants in the UKB (n=490,413) **(Figure 3A).** For the 12 *RB1* pathogenic variant carriers in UKB, the age-related penetrance was lower than expected based on estimates previously published in literature. For *RB1* variant carriers, the penetrance by age 18 was 25.0% (95% CI 0-45.9) and remained at this value by 60 years of age using the stringent Rb phenotype (**Figure 3A**). Relaxing the phenotype criteria to include any ocular and Rb-associated non-ocular cancers (permissive phenotype), the penetrance started at 41.7% (95% CI 0.59-63.8) by age 18 and increased to 50.0% (95% CI 12.0-71.6) by 60 years of age. Overall population ascertained *RB1* variant carriers exhibit a reduced penetrance in the childhood period, which is most relevant to newborn screening.

**Figure 3:**
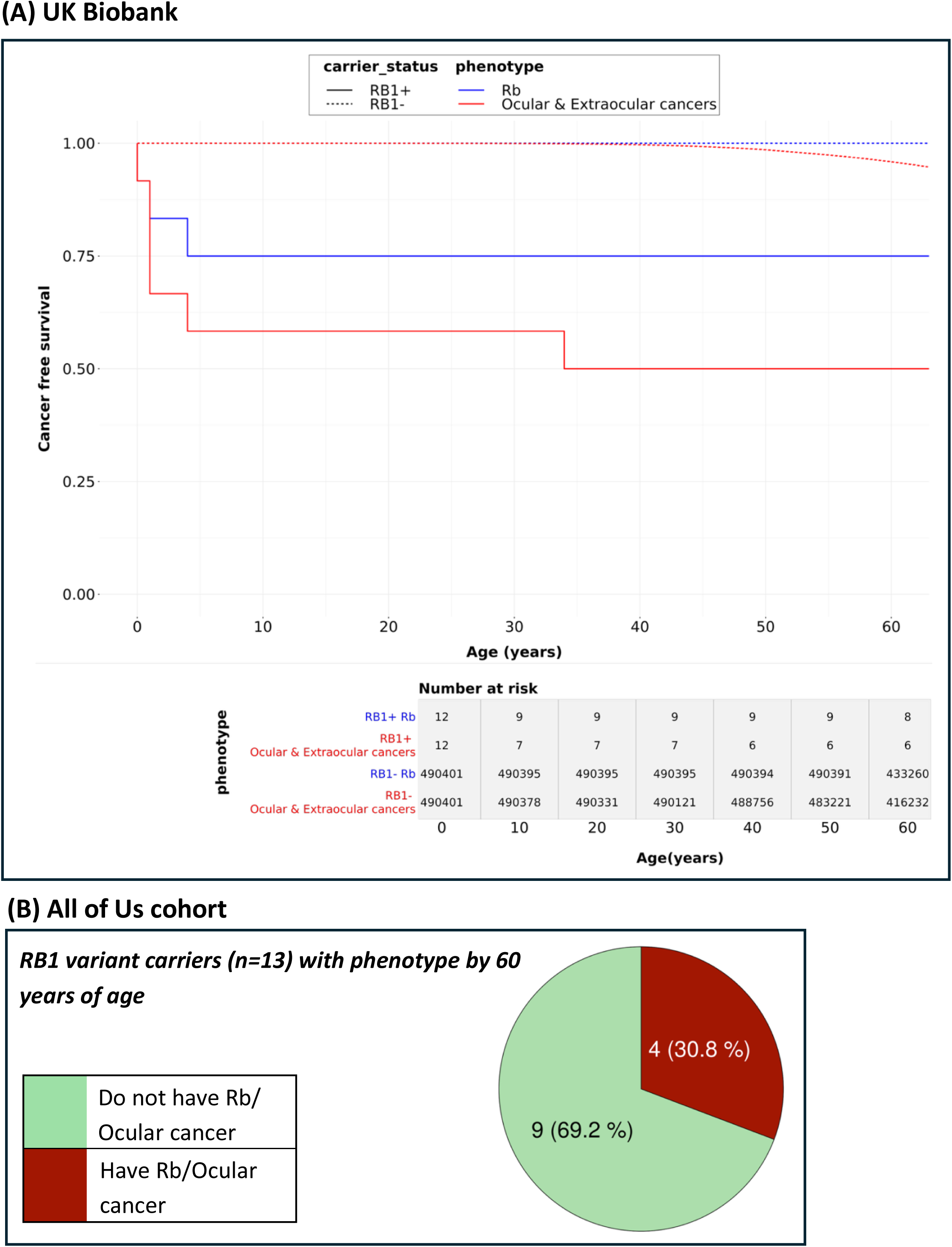
Penetrance of *RB1* variants and retinoblastoma in the UK Biobank (UKB) and All of Us (AoU) cohorts. Figures depicting the proportion of RB1 variant carriers who develop a phenotype consistent with heritable retinoblastoma (hRb). Figure (A) depicts the Kaplan-Meier survival probability (proportion of individuals that remain cancer free) is shown on the Y axis and the age at risk is shown on the X axis. Survival curves were calculated for *RB1* variant carriers (n=12) and non-carriers (n=490,401) and were plotted for the stringent (blue) phenotype criteria, consistent with a diagnosis of retinoblastoma (Rb) and the permissive (red) phenotype criteria, which included any ocular and Rb-associated extraocular cancers. Age-related penetrance estimates were calculated using this survival model, specifically 1 - [Kaplan-Meier survival probability at age X]. Figure (B) depicts the proportion of variant carriers with diagnostic codes for any ocular cancers in AoU. Due to the lack of a precise age of diagnosis in self-reported medical records, we were unable to perform an equivalent Kaplan-Meier survival analysis in this cohort.

### Carriers of *RB1* pathogenic variants in the All of Us cohort exhibit reduced penetrance compared to previously reported estimates

Of the 317,964 AoU participants with both WGS and EHR data available we identified 13 high-confidence (likely) pathogenic or pLoF *RB1* variants in 13 individuals, two of which had also been identified in the UKB cohort (**Table 1**, **Figure 2**). One participant carried two out-of-frame insertions in the *RB1* gene (c.743_744insGTATAACCCATTAATGGGTATAATG (p.Ser249TyrfsTer30) and c.746_747insTG (p.Pro250AspfsTer15)) that together caused an in-frame insertion that introduced a premature stop codon, and so was included in our final variants list. Similar to the UKB, most variants had also previously been submitted to ClinVar and classified as P/LP (**Table 1**). Due to the lack of cancer registry data in this cohort, we screened participants for features consistent with hRb using a broader set of diagnostic codes, which would include participants with any ocular cancers (Rb and/or ocular cancer, n=295). Despite using a broader set of diagnostic codes to define hRb, Fisher’s exact test revealed a significant enrichment of ocular cancers in *RB1* pathogenic variant carriers, p=5.16x10^-10^ (OR: 485.5) compared to non-carriers in this cohort. Only four of the 13 *RB1* variant carriers had diagnostic codes or self-reported Rb and/or ocular cancer by 60 years of age, which equates to a penetrance of 30.8% (4/13) **(Figure 3B).** This is consistent with the reduced penetrance observed in the UKB, despite being a completely different adult population cohort. Although the age of diagnosis in AoU is available through EHRs, self-reported records do not provide a specific age of diagnosis (see methods). We were therefore unable to perform an equivalent survival analysis in AoU.

### The calculated prevalence of Rb suggests that adult population cohorts show no evidence of depletion for affected individuals

Based on the reported lifetime prevalence of Rb, estimated between 1-9 in 100,000 (7), we can calculate the expected prevalence of Rb in UKB and AoU. Participants who met the stringent phenotype were used to estimate the prevalence of Rb in the UKB. Due to the lack of cancer registry data in AoU, we took a broader approach when classifying an Rb phenotype compared to UKB to ensure we captured all possibly affected individuals. To estimate Rb prevalence in AoU, we used SNOMED CT codes specifically corresponding to malignant neoplasm of the retina **(Table S2).** We compared the number of individuals with Rb in the UKB (n=21) and AoU (n=28) against the expected number of cases to determine the extent of depletion in these cohorts. For each cohort, the total number of individuals with Rb was within expected ranges according to the calculated upper and lower bounds (UKB: 5-44 & AoU: 3-29) **(Figure 4).** This remained consistent upon combining both cohorts (n=808,377, bounds: 8-73) **(Figure 4).**

**Figure 4:**
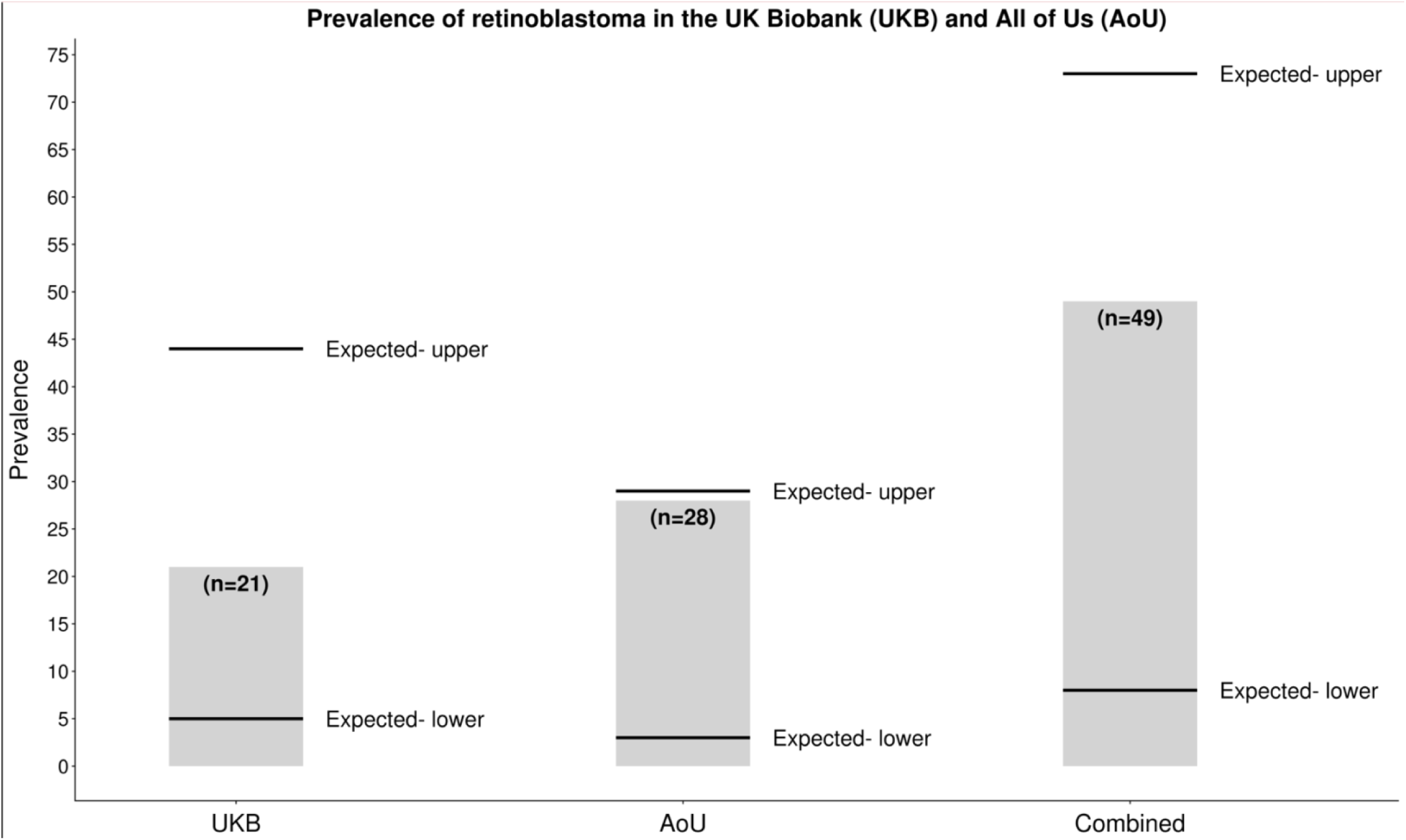
Prevalence of retinoblastoma in the UK Biobank (UKB) and All of Us (AoU) cohorts. Observed prevalence in each cohort is shown by the grey bar. The upper and lower number of expected cases are highlighted by the black lines. Expected prevalence was calculated using the estimated lifetime prevalence of retinoblastoma of 1-9 in 100,000 for UKB (n=490,413), AoU (n=317,964) and combined cohorts (n= 808,377)

In the UKB, only 14.3% (3/21) individuals with Rb also carried a pathogenic *RB1* variant, despite detecting 12 pathogenic variant carriers in the cohort. Similarly, only a small proportion of variant carriers in AoU overlapped with those who had Rb-related diagnostic codes (10.7%, 3/28). A proportion of Rb-positive individuals will represent non-heritable Rb cases caused by somatic *RB1* mutations in the retina (15). Highly penetrant *RB1* variants may be less common in these cohorts, which could explain the minimal overlap between *RB1* variant carriers and those with Rb-related diagnostic codes. In the UKB we were able to assess these individuals for deletions encompassing the *RB1* gene, however there were no copy number variants detected in these individuals. It is also possible that we have not detected *RB1* pathogenic variants due to poor coverage of WGS data or due to low-level mosaic cases where the variant was undetectable in blood.

## Discussion

The association between *RB1* and Rb is well-established, yet there is still limited understanding of the penetrance of *RB1* pathogenic variants in the general population. We screened two adult population-based cohorts, the UKB (n=490,413) and AoU (n=317,964), for *RB1* pathogenic variants to estimate the penetrance and prevalence of hRb. We have shown that, in both cohorts, *RB1* penetrance was substantially lower compared to the >90% penetrance reported in literature (7,8,18). Combined meta-analysis revealed a 28.0% (7/25) penetrance for *RB1* variant carriers across both the UKB and AoU cohorts. The prevalence of Rb was consistent with the expected number of cases for each cohort, suggesting minimal depletion of Rb. This result is consistent with other studies showing reduced penetrance of a variety of conditions in clinically unselected populations (38–40), and has major implications for individuals detected through a genotype-first screening approach.

All 22 *RB1* variants included in this study were annotated as high impact, predicted to significantly impact protein function; the majority had also been classified as P/LP in ClinVar. Of these, 19 *RB1* variants were predicted to result in complete loss of the protein, previously associated with a high penetrance (8,19,20). Only three variants have reports of a reduced penetrance in family cohort studies, all of which have been classified as P/LP in ClinVar at a 1 or 2* level. This included two splicing variants (c.539+1G>A, c.862-1G>A), predicted to lead to in-frame exon skipping, which is a recognised mechanism for reduced penetrance (19,20). Despite leading to out-of-frame exon skipping, one splice-variant (c.607+1G>T) has been associated with a variable penetrance in family cohort studies, possibly explained by additional genetic modifiers influencing *RB1* expression (21,41). Recently, published guidelines on the variant prioritisation criteria that will be used in the UK’s gNBS programme – the Generation Study (42) – included variants classified as P/LP in the ClinVar database, with no more than one Benign/Likely Benign classification, and high-impact variants predicted to severely disrupt protein function (42). Our approach was therefore consistent with guidelines used by existing gNBS programmes, and in some cases took a more stringent approach to variant curation (42). All *RB1* variants included in this study would be prioritised and considered clinically actionable according to these criteria.

The number of Rb diagnoses in the UKB and AoU was consistent with the expected number of cases for each cohort, based on the reported lifetime prevalence of Rb (7), which is in keeping with the high 5-year survival rates for individuals diagnosed with Rb (13). However, the minimal overlap between *RB1* variant carriers and those with Rb-related diagnostic codes, could indicate a depletion of highly penetrant *RB1* pathogenic variant carriers in these cohorts. In the UKB incidental findings detected through retinal imaging data would be reported back to participants (43), so it would be unlikely that atypical late Rb diagnoses were missed. The UKB is an older population cohort (mean age of 57 years at recruitment), and biased towards healthier individuals (44). This can introduce a survival bias, where those with severe disease may have died prior to enrolment or be too unwell to participate, which could also contribute to the reduced penetrance in this cohort. In contrast, AoU participants appear to have a lower general health compared with the general US population (45). Despite this, *RB1* pathogenic variant carriers still showed reduced penetrance, which was remarkably consistent across both cohorts, and limited overlap between individuals diagnosed with Rb.

In general, a risk of ≥5% of developing a paediatric cancer by 20 years of age is enough to warrant surveillance measures (46). For Rb specifically, surveillance is recommended for anything above the general population risk of 0.007% (15). Therefore, despite the reduced penetrance seen in this study, it is still high enough to consider newborn screening, due to the benefits of early detection to improve survival and vision outcomes (47,48). Moreover, gNBS programmes provide the opportunity to identify newborns with *de novo RB1* variants, which up until now have only been detected upon clinical presentation of Rb (15,16). The lower penetrance estimates observed across UKB and AoU provide new data that may be more appropriate than estimates from clinical cohorts when counselling families where an *RB1* variant is identified in the context of gNBS with no existing family history of Rb. Whilst for some cancer predisposition syndromes, lower penetrance estimates may result in modification of cancer surveillance or risk reducing interventions, the observed lower penetrance for *RB1* in this study still falls at a threshold at which Rb surveillance is indicated based on current clinical recommendations given that direct visualisation of the retina provides a unique opportunity to detect and treat tumours at an early stage to minimise loss of vision and/or eyes.

Current guidelines for individuals with diagnostically confirmed *RB1* variants recommend an intensive screening protocol, which involve regular hospital visits and EUAs (9,15,16). These have been developed based on results from clinically ascertained cohorts (15,16), and most studies completed on pre-symptomatic screening of at-risk children have included those who are already affected or with a known family history of the condition (47). These studies have not considered the implications of Rb screening in individuals with *RB1* pathogenic variants detected using genotype-first screening approaches such as gNBS programmes, most of whom will have no family history or symptoms at the point of testing. Potential harms of repeated exposure to anaesthetic medicines are recognised (49). For those at high risk of developing Rb, the benefits of early detection are considered to outweigh this potential harm. However, as more results are published from gNBS and additional population-based studies are completed, if this reduced penetrance is further established and quantified, it may be appropriate to review surveillance protocols for *RB1*-positive individuals ascertained through population-based testing in consultation with the appropriate clinical experts and patient groups.

Our study had several limitations. Firstly, penetrance and prevalence estimates were dependent on the availability and accuracy of clinical coding and responses provided in linked healthcare data and self-reported medical conditions in the respective biobanks. Family history of eye cancer was unavailable in UKB. Although this was recorded in AoU, none of the responding *RB1* variant carriers reported a history of eye cancer. Clinical coding is not always complete and is often allocated to determine healthcare costs (50). In both cohorts we used an expanded set of clinical codes for our penetrance estimates that could account for incorrect allocations. Future work could assess the accuracy of diagnostic code allocations to further validate these records, as done in other studies (50).

Secondly, neither cohort is representative of the general population of newborns that might be offered screening. The UKB is known to suffer from healthy-volunteer recruitment biases (44). Estimates generated from this cohort therefore likely represent lower bounds of the penetrance (4,44). The penetrance of *RB1* pathogenic variants most representative of the general population likely lies within the proposed estimates of 25.0-50.0%. Nonetheless, the presence of *RB1* pathogenic variants in adults >40 years old who were healthy enough to volunteer to be part of a population cohort indicates that penetrance is incomplete.

Thirdly, heritable Rb is a rare genetic condition and despite the large cohorts, only a small proportion of UKB and AoU participants had an *RB1* variant after curation. This rarity also prevented comparison of the penetrance between individual variants. Larger cohorts are required to improve understanding of individual variant penetrance.

## Conclusion

This study combined genotype- and phenotype-first approaches to gain a better understanding of the prevalence and penetrance of hRb in two adult population cohorts. The penetrance of high impact pLoF *RB1* variants was between 25.0-50.0% across the UKB and AoU cohorts, with a combined penetrance of 28.0%. This result has important implications for future genotype-first approaches, as we have shown that a large proportion of variant carriers may not develop the condition. Although screening for Rb facilitates early detection and optimal prognosis, we have shown that the risk of developing the condition is lower in a population setting than a standard diagnostic context. Careful consideration will be required when communicating the risk of *RB1* variants to families of newborns detected through gNBS programmes.

## Data and code availability statement

Data from UK Biobank and All of Us cohorts cannot be shared publicly because of data availability and data return policies. Data are available for researchers who meet the criteria for access to datasets to UK Biobank (www.ukbiobank.ac.uk) and All of Us (https://allofus.nih.gov).

Code used to extract data from the UK Biobank Healthcare Records are available at https://github.com/hdg204/UKBB/tree/main. Code used for analysing data and generating plots is available at https://github.com/ILazaridi/RB1_penetrance

## Acknowledgments and funding statements

We would like to thank Dr Alison Foster for their expert clinical input on the manuscript.

We gratefully acknowledge participants of the UK Biobank and All of Us for their contributions, without whom this research would not have been possible. This research has been conducted using the UK Biobank Resource under Application Number 103356 and uses data provided by patients and collected by the NHS as part of their care and support. We also thank the National Institutes of Health’s All of Us Research Program for making available the participant data examined in this study. The authors would like to acknowledge the use of the University of Exeter High-Performance Computing (HPC) facility in carrying out this work.

This work was supported by the Medical Research Council [MR/X021351/1] and the National Institute for Health and Care Research Exeter Biomedical Research Centre. The views expressed are those of the author(s) and not necessarily those of the NIHR or the Department of Health and Social Care.

## Author Contribution Statement

I-AL- Formal analysis, Investigation, Visualisation, Writing- original draft, Writing – review & editing

TH- Methodology, Writing- review & editing

HH- Writing – review & editing

JF- Writing – review & editing

ELB- Funding curation, Writing – review & editing

MNW- Funding curation, Data curation, Writing – review & editing

CFW- Funding curation, Conceptualisation, Writing- original draft, Writing – review & editing

LJ- Funding curation, Conceptualisation, Data Curation, Supervision, Writing- original draft, Writing – review & editing

## Ethical Approval

UK Biobank protocols were approved by the National Research Ethics Service Committee. This study was performed in a manner that aligned with the ethical principles set by All of Us-Policy on the Ethical Conduct of Research.

## Competing Interests

The authors declare no competing interests.

## Notes

### Competing Interest Statement

The authors have declared no competing interest.

### Author Declarations

This study used anonymized data from UK Biobank under approved project application 103356. Research Tissue Bank status granted to UK Biobank by the North West Multi-centre Research Ethics Committee of the Health Research Authority (reference 16/NW/0274) and was renewed in 2021. The All of Us Institutional Review Board provided ethical approval of all protocols and materials used by the All of Us Research Programme. All participants provided informed consent before participating in the All of Us programme.

