## Supplemental data for "Prevalence and Penetrance of Heritable Retinoblastoma in Two Adult Population Cohorts: Implications for genomic newborn screening"

Associated phenotypes mapped to the relevant international classification of disease (ICD-9/10), Office of Population Censuses and Surveys codes (OPCS-4) and SNOMED CT codes. Clinical features associated with RB1-hereditary RB were identified using Gene2Phenotype (ID: G2P01817) and relevant cohort studies (1–10). These were discussed with a wider team of clinicians and clinical geneticists.

**Table 1: Diagnostic codes used in stringent and permissive phenotype criteria in the UK Biobank cohort**

**STRINGENT CRITERIA**

| Self-reported records | | | |
|---|---|---|---|
| ***Code*** | ***Description*** | | |
| X00eS | Retinoblastoma | | |
| **HES and/or cancer registry records** | | | |
| ***ICD10*** | ***Description*** | ***ICD-9*** | ***Description*** |
| C69.2 | Malignant neoplasm of eye and adnexa; Retina | 1905 | Malignant neoplasm of retina |

**PERMISSIVE CRITERIA***

| Self-reported records | | | |
|---|---|---|---|
| ***Code*** | ***Description*** | | |
| X00eS | Retinoblastoma | | |
| 72001 | Eye and/or adnexal cancer | | |
| **HES and/or Cancer registry records** | | | |
| ***OPCS-4*** | ***Description (OPCS-4)*** | | |
| C01 | Excision of eye | | |
| C91 | Operations on anterior segment of eye | | |
| F29 | Correction of deformity of palate | | |
| ***ICD-10*** | ***Description (ICD-10)*** | ***ICD-9*** | ***Description (ICD-9)*** |
| C69 | Malignant neoplasm of eye and adnexa | 190 | Malignant neoplasm of the eye |
| C69.2 | Malignant neoplasm of eye and adnexa; Retina | 1905 | Malignant neoplasm of the retina |
| C75.3 | Malignant neoplasm of pineal gland | 1944 | Malignant neoplasm of pineal gland |
| Q35 | Cleft palate | 749 | Cleft palate and cleft lip |
| C34 | Malignant neoplasm of bronchus/lung | 162 | Malignant neoplasm of trachea/bronchus/lung |
| C40-41 | Malignant neoplasms of bone and articular cartilage | 170 | Malignant neoplasm of bone and articular cartilage |
| C45-49 | Malignant neoplasms of mesothelial and soft tissue | 171 | Malignant neoplasm of connective/other soft tissue |
| C43 | Malignant melanoma of skin | 172 | Malignant melanoma of skin |
| C50 | Malignant neoplasm of the breast | 174/175 | Malignant neoplasm of female/male breast |
| C67 | Malignant neoplasm of bladder | 188 | Malignant neoplasm of bladder |
| C71 | Malignant neoplasm of brain | 191 | Malignant neoplasm of brain |
| C72 | Malignant neoplasm of spinal cord, cranial nerves and other parts of central nervous system | 192 | Malignant neoplasm of other and unspecified parts of nervous system |
| C91-95 | Lymphoid, Myeloid, Monocytic and other leukaemias of specified/unspecified cell types | 204-208 | Lymphoid, Myeloid and Monocytic and other leukaemias of specified/unspecified cell types |
| H33 | Retinal detachment and breaks | 361 | Retinal detachment |

ICD-9, ICD-10, OPCS-4 and self-reported codes and associated descriptions screened for in the Hospital Episode Statistic (HES), cancer registry and self-reported records. All associated subcodes were included unless specified.

** This criteria includes all codes used in the stringent phenotype category*

**Table 2: Diagnostic codes used in the All of Us cohort**

| Phenotype criteria | |
|---|---|
| **Personal and Family Health History survey** | |
| ***Code*** | ***Description*** |
| - | Including yourself who in your family has had eye cancer? -Self |
| **Electronic Health records** | |
| ***SNOMED*** | ***Description*** |
| **371986009** | **Primary malignant neoplasm of eye (group)** |
| 109948008 | Overlapping malignant neoplasm of eye and adnexa (primary) |
| 93764002 | Primary malignant neoplasm of conjunctiva of eye |
| 763477007 | Primary lymphoma of conjunctiva |
| 231833004 | Sebaceous adenocarcinoma of eyelid |
| 93766000 | Primary malignant neoplasm of cornea of eye |
| 93987004 | Primary malignant neoplasm of retina |
| 735916009 | Primary malignant neuroepithelial neoplasm of retina |
| 370967009 | Retinoblastoma |
| 94128004 | Primary malignant neoplasm of uveal tract of eye |
| 93755007 | Primary malignant neoplasm of choroid |
| 93756008 | Primary malignant neoplasm of ciliary body (primary) |
| **363465007** | **Malignant tumour of retina (group)** |
| 93987004 | Primary malignant neoplasm of retina |
| 735916009 | Primary malignant neuroepithelial neoplasm of retina |
| 370967009 | Retinoblastoma |
